# Home-based virtual reality scenic relaxation for patients with insomnia and anxiety: a single-arm feasibility and safety study

**DOI:** 10.1101/2025.11.06.25339711

**Authors:** Nishiki Yanagi, Narimasa Katsuta, Shuko Nojiri, Tamaki Nara, Kanako Kumamaru, Yoshihide Takeshita, Morikuni Tobita

## Abstract

Insomnia commonly co-occurs with anxiety, and pre-sleep arousal may sustain both conditions. Virtual reality-based relaxation can help to downregulate arousal before sleep. There is limited access to cognitive behavioral therapy for insomnia, and few studies have examined VR relaxation in routine clinical practice. The study aimed to evaluate the feasibility and safety of a 4-week home-based virtual reality scenic relaxation intervention for outpatients with chronic insomnia and anxiety symptoms, and to explore changes in anxiety, depression, and sleep. We conducted a single-arm, pre–post feasibility study at an outpatient clinic. Adults with chronic insomnia and co-occurring clinically significant anxiety symptoms (Generalized Anxiety Disorder-7 [GAD-7] ≥5 and Pittsburgh Sleep Quality Index [PSQI] ≥6) completed eight home virtual reality sessions (approximately 20 min each) over 4 weeks. Primary outcomes were feasibility (recruitment, retention, and adherence) and safety (virtual reality-related adverse events). The exploratory outcomes were changes in the GAD-7, Patient Health Questionnaire-9 [PHQ-9], and PSQI scores from baseline to 4 weeks. The analyses were primarily descriptive, with effect sizes and 95% confidence intervals reported. Of the 11 enrolled participants, 10 completed the study (retention 91%) and achieved 8/8 sessions. No serious adverse events occurred, and only mild, transient discomfort was reported by 3 of 10 participants (30%). The participants reported moderate-to-high levels of satisfaction with their experience and a moderate-to-high sense of immersion in the virtual reality environment. In exploratory analyses, mean (standard deviation) changes were −4.0 (2.8), −4.1 (4.5), and −2.0 (2.1) for GAD-7, PHQ-9, and PSQI, respectively. Most participants improved on ≥1 outcome. Home-based virtual reality scenic relaxation is feasible, acceptable, and safe for participants with insomnia and anxiety, with exploratory indications of symptom improvement. Controlled trials are warranted to establish efficacy and inform optimal implementation.

**Trial registration:** Japan Registry of Clinical Trials (https://jrct.niph.go.jp/; registry number: jRCT1032240289; date of registration: August 22, 2024).

## Introduction

Chronic insomnia is common and burdensome, affecting a substantial proportion of adults and frequently co-occurring with anxiety symptoms. In Japan, a nationwide interview survey estimated that approximately 12–15% of adults meet criteria for insomnia, underscoring its public health impact [1,2]. Sleep difficulties are associated with daytime impairment, reduced quality of life, and increased healthcare use.

Cognitive behavioral therapy for insomnia (CBT-I) is the first-line non-pharmacological treatment for chronic insomnia, with strong evidence for improving sleep onset, maintenance, and overall sleep quality [3]. In routine care, however, CBT-I can be unavailable or unsuccessful; in such cases, short-term pharmacotherapy may be considered through shared decision-making [3]. These gaps motivate the development of scalable, acceptable, and engaging adjuncts that reduce pre-sleep arousal and are particularly suitable for patients who report anxiety symptoms.

Immersive virtual reality (VR) is one such adjunct because it can be delivered at home on consumer headsets and directly targets arousal through controlled, engaging environments. Traditionally, VR therapy has been used to treat phobias and anxiety disorders by simulating situations that elicit negative emotional experiences (including virtual airplane rides for individuals with aviophobia) [4]. However, recent research has focused on immersive VR experiences that elicit positive emotions and facilitate relaxation and mindfulness [5,6]. This approach aims to help patients focus on positive stimuli and leverage these states to improve their quality of life.

VR therapy offers a unique way to deliver immersive relaxation and mindfulness. By transporting users into calm virtual environments, VR induces relaxation and reduces anxiety in experimental and clinical settings [7,8]. For example, the VR mind–body approach downregulates psychophysiological arousal in adolescents with insomnia [9]. VR meditation improves stress and anxiety in younger adults [10]; its effects on older adults are under investigation [11].

Notably, VR-based interventions are being explored across various patient groups. Lai et al. reported that a 4-week home-based group VR gaming program for adolescents with physical disabilities had good acceptability, with participants reporting high enjoyment and social connection. However, retention was 67% [12]. For frontline healthcare workers during the coronavirus disease 2019 pandemic, a protocol by Pallavicini et al. is testing a brief home VR relaxation training (“MIND-VR” with a nature-based scene) to manage stress and anxiety using the Meta Quest 2 [13]. Similarly, an upcoming randomized trial in older adults will evaluate the feasibility and effects of VR-guided meditation (eight sessions over 4 weeks) on stress, anxiety (Generalized Anxiety Disorder-7 [GAD-7]), depression (Patient Health Questionnaire-9 [PHQ-9]), and insomnia severity [11]. Collectively, these studies indicate feasibility of VR in mental health and sleep medicine.

Despite encouraging early data on VR for relaxation, most studies have been short-term, often in healthy volunteers, and very few have specifically examined insomnia outcomes in patients with anxiety. We conducted a single-arm feasibility study of a 4-week home-based VR scenic relaxation intervention to evaluate safety, acceptability, adherence, and exploratory pre-to-post changes in anxiety, depression, and sleep. This study serves as an initial step in informing future controlled trials of VR as a complementary therapy for this population.

## Materials and Methods

### Study Design and Setting

This single-arm, pre–post feasibility study was conducted at the outpatient mental health clinic of Juntendo University Hospital (Japan). Adults were screened between October 2024 and April 2025 and received VR intervention in addition to usual care.

### Recruitment and Eligibility

Potential participants were approached during routine visits and prescreened by treating clinicians. Inclusion criteria: (1) Age 18–59 years; (2) co-occurring anxiety symptoms and chronic insomnia, irrespective of the primary Diagnostic and Statistical Manual of Mental Disorders (DSM-5) diagnosis (for example, participants could have an anxiety disorder or another psychiatric condition characterized by prominent anxiety features); (3) GAD-7 ≥5 [14]; (4) Pittsburgh Sleep Quality Index [PSQI] ≥6 [15]; and (5) ability and willingness to use a home VR headset (confirmed during consent and brief device orientation).

The exclusion criteria were as follows: Use of VR deemed clinically unsafe (including uncontrolled epilepsy, severe vestibular disorder), pregnancy or possible pregnancy, severe uncorrected visual or hearing impairment or other physical limitations preventing safe use, lack of required home Wi-Fi, any condition judged by the investigator to make participation inappropriate (including acute psychosis), or screening failure on symptom thresholds (GAD-7<5 or PSQI<6).

Participants were prescreened between October 2024 and April 2025. Enrollment occurred from December 6, 2024 through April 3, 2025. All eligible individuals provided written informed consent before any study procedures. The protocol was approved by Juntendo University Certified Review Board (J24-007) and conducted in accordance with the Declaration of Helsinki. Trial registration: Japan Registry of Clinical Trials (jRCT1032240289), registered prior to enrollment.

### VR Relaxation Intervention

Participants completed a 4-week [11], home-based [13,16] VR scenic relaxation intervention. The intervention comprised eight sessions [11] (sessions were spaced by ≥48 h to reduce potential after-effects, a pragmatic choice informed by cybersickness literature [17]) delivered on a standalone Meta Quest 2 headset (Fig 2A). VR content was limited to two scenes provided under “VR Relaxation,” viewed in a fixed sequence within each session: (i) Maldives ocean panorama (Fig 2B) followed by (ii) New Zealand Lake Tekapo panorama (Fig 2C). Each session lasted approximately 20 min (two approximately 10-minute scenes back-to-back) with ambient audio and paced-breathing prompts [9]. The participants were instructed to sit comfortably, minimize head/body movements to reduce the risk of cybersickness, and stop if discomfort occurred. The sessions were self-scheduled (preferably approximately 2 h before going to bed) to facilitate relaxation and reduce arousal before sleep or during periods of anxiety.

**Fig 1.**
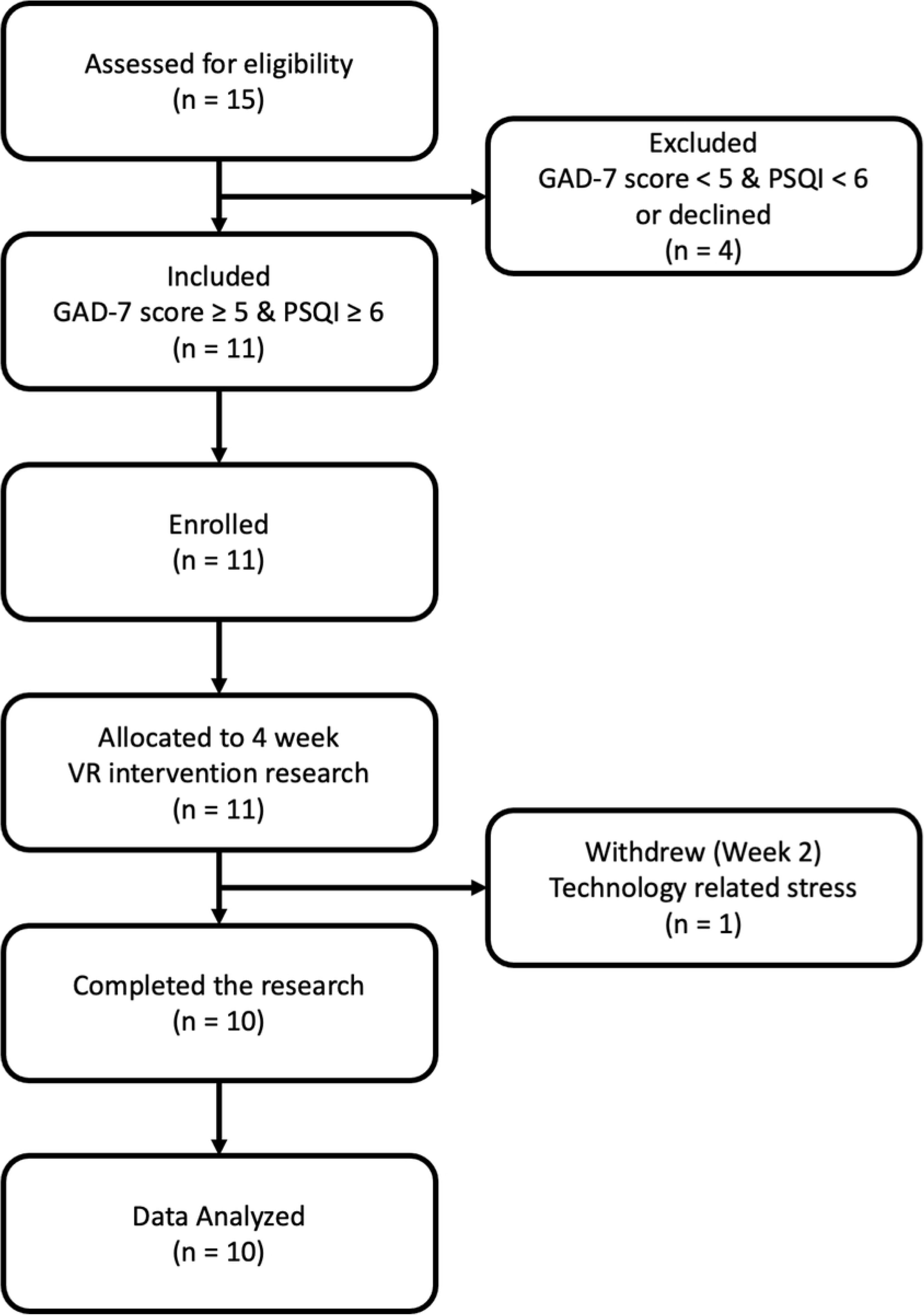
Participant flow. Eleven participants met eligibility criteria (GAD-7 ≥5; PSQI ≥6) and were enrolled. One withdrew at Week 2 due to technology-related stress. Ten completed the intervention and were included in the analysis. Abbreviations: GAD-7, Generalized Anxiety Disorder-7; PSQI, Pittsburgh Sleep Quality Index.

**Fig 2.**
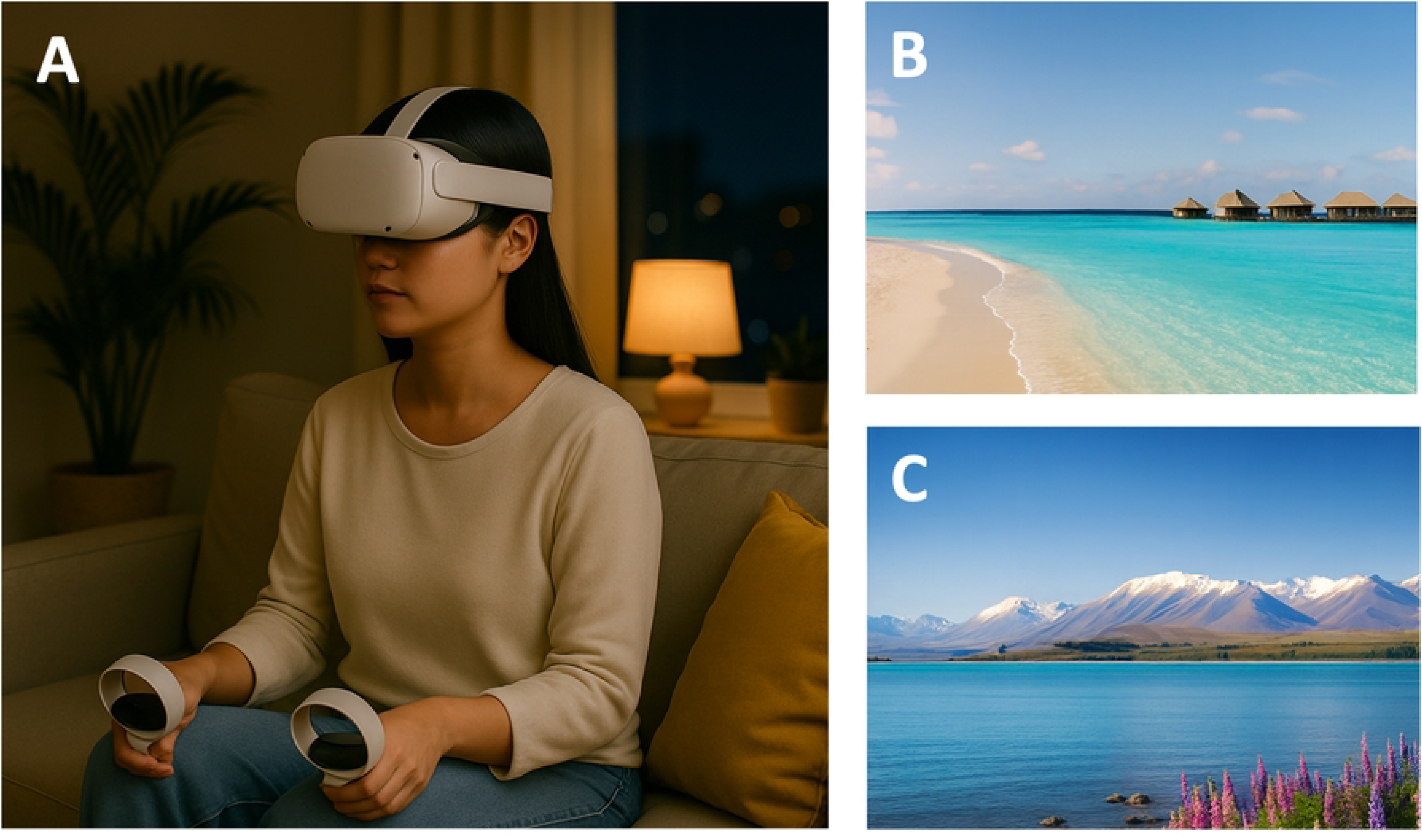
Standalone VR headset posture and example scenic relaxation environments (illustrative). (A) Seated home-use posture with a standalone virtual reality (VR) headset and two hand controllers. (B) Ocean panorama (“Maldives” motif). (C) Lake panorama (“Lake Tekapo” motif). Images are illustrative only and do not depict actual app assets or participant images. AI disclosure: Panels A, B, and C were generated by the authors in January 2025 using an AI image model from OpenAI. We used illustrations because reproduction of in-app screenshots and product images is restricted by license. No AI-generated images were analyzed, and no clinical inference relies on these illustrations.

### Procedures

We implemented a remote home-based delivery model (Fig 3). Candidates were pre-identified by the attending physician during routine care and invited to a clinic visit (screening and onboarding; Fig 3, Steps 1–2). During this visit, the participants first completed screening questionnaires (GAD-7, PSQI, and PHQ-9, if applicable). The investigators reviewed eligibility according to the inclusion and exclusion criteria. All eligible individuals provided written informed consent. After consent was obtained, demographics and baseline questionnaires (GAD-7, PHQ-9, and PSQI) were recorded. If the screening questionnaires had just been completed, the scores were documented as baseline. The participants then received a device orientation, a brief hands-on trial, and verification of their access to the assigned VR content.

**Fig 3.**
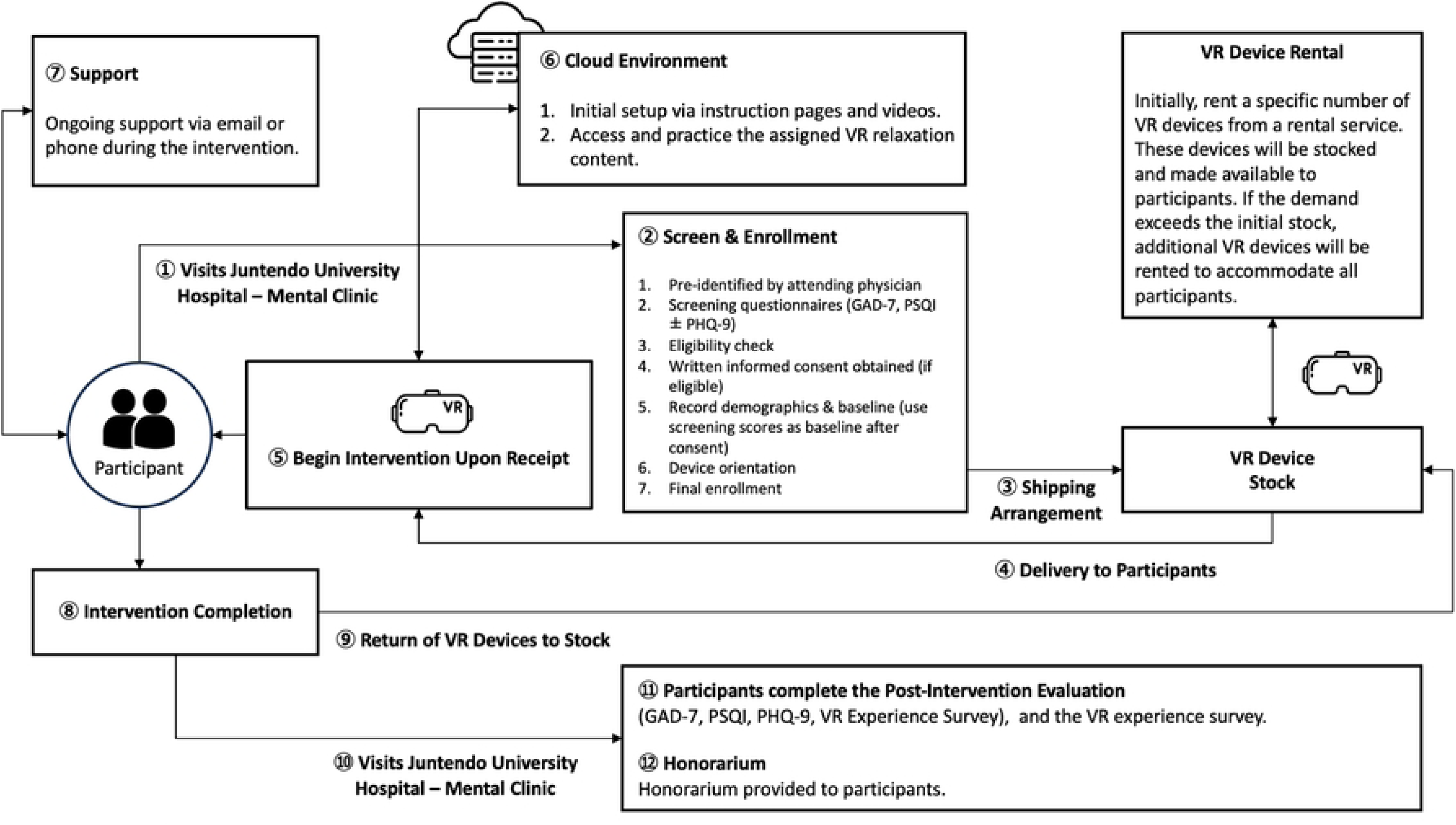
Study process flowchart. VR Headsets (Meta Quest 2) were rented and shipped to participants; eight home-based VR sessions were completed over 4 weeks with remote support as needed; devices were returned, and post-intervention evaluations were conducted at the clinic.

The VR headsets were provided by a rental service and prepared by the team. Shipping was arranged (Fig 3, Step 3), and the devices were delivered to the participants (Fig 3, Step 4). On receipt, the participants began the 4-week home intervention (Fig 3, Step 5) with a target of two sessions per week (eight total). Setup resources were available in the cloud environment (Fig 3, Step 6), and support was available via email or phone during the intervention (Fig 3, Step 7).

After completion of the eight sessions or at 4 weeks (Fig 3, Step 8), devices were returned to stock by courier or at the post-intervention visit (Fig 3, Steps 9–10). During the clinic visit (Fig 3, Step 10), the staff performed a brief device check and a debriefing. The participants then completed the post-intervention evaluation (Fig 3, Step 11)— the GAD-7, PHQ-9, PSQI, and a VR experience survey—and received an honorarium (Fig 3, Step 12).

Evaluations were scheduled before (baseline, week 0) and after (after the 4-week intervention). An interim assessment was not performed. Adherence was monitored using participant session date logs and brief weekly email or phone check-ins. Deviations were queried and corrected, and device receipt prior to baseline and return after the intervention were confirmed by email or phone.

### Outcome Measures

Feasibility and Engagement: We tracked recruitment, retention (proportion completing the 4-week intervention and assessments), and adherence (≥75% of sessions completed considered adequate). The reasons for attrition or missing sessions were also recorded. Post-intervention, the participants completed a brief VR experience survey (five Likert-type items: ease of use, immersion, content quality, comfort, and willingness to use again), which was summarized descriptively.

Clinical outcomes (exploratory): The primary clinical outcomes were changes in anxiety, insomnia, and related symptom severities from baseline to post-intervention. Anxiety was assessed with the GAD-7 (0–21). Because insomnia is characterized by persistently poor sleep quality, curtailed or fragmented sleep, and daytime dysfunction, we used the PSQI [15] to quantify sleep quality as an index of insomnia-related impairment. We analyzed the PSQI global score (0–21; >5 denotes poor sleep quality) and its seven components (0–3 each; higher = worse): C1 subjective sleep quality; C2 sleep latency; C3 sleep duration; C4 habitual sleep efficiency; C5 sleep disturbances; C6 use of sleeping medication; C7 daytime dysfunction.

Component-level results are summarized in S1 Table. The secondary outcome was depression symptoms measured using the PHQ-9 (score, 0–27) [18]. These validated self-report scales are commonly used in anxiety and insomnia research. Higher scores indicated worse symptoms. All questionnaires were completed at baseline and post-intervention.

Safety monitoring: We assessed adverse events (AEs) at each contact and post-intervention visit. An AE was defined as any untoward medical occurrence temporally associated with VR use, whether it was causally related [19]. Serious AEs (SAEs) were defined as events that resulted in death, were life-threatening, required inpatient hospitalization or prolongation of hospitalization, resulted in persistent or significant disability or incapacity, involved a congenital anomaly or birth defect, or were judged as important medical events requiring intervention to prevent one of these outcomes [19]. Investigators graded AE severity as mild, moderate, or severe and relatedness as unrelated, possibly related, or probably related to VR. The expected VR-related AEs included dizziness, nausea, eye strain, and headache [17,20]. No SAEs occurred.

### Data Analysis

Given the small sample size and exploratory nature of the study, the analyses were descriptive. We did not conduct null hypothesis significance testing, and p-values are not reported. For continuous outcomes (GAD-7, PHQ-9, and PSQI), we summarized the pre- and post-intervention values as means and standard deviations (SD), medians with interquartile ranges, and ranges, as well as the within-person mean change (post−pre) with 95% confidence intervals (CI). To aid interpretation, we provided standardized within-subject effect sizes (Cohen’s dz) and approximated 95% CIs. Categorical summaries (including severity bands) were presented as counts and percentages (95% CIs, where informative). The feasibility metrics (recruitment, retention, and adherence) are shown as proportions (with binomial CIs when appropriate). The VR experience survey items were summarized using item-level distributions and descriptive statistics. Missing outcome data were not imputed. Analyses used available paired observations and were interpreted as exploratory without adjustment for multiple comparisons.

As a feasibility pilot, we targeted a pragmatic sample size of approximately 10 participants to estimate adherence and safety parameters, consistent with recommendations for pilot studies [21–23].

This study was reported in accordance with the Transparent Reporting of Evaluations with Nonrandomized Designs (TREND) statement [24]; see S1 Checklist.

## Results

### Participant Flow and Feasibility

We approached 15 potential participants who were referred by their primary care physicians. Of these, one patient declined participation owing to scheduling conflicts, and three did not meet the eligibility criteria during screening. Ultimately, 11 patients passed the screening and signed the consent forms. Fig 1 shows the participant flow. One participant withdrew from the study during week 2, citing increased stress and frustration while operating the VR headset. The participant had a history of bipolar disorder and reported feeling “overwhelmed” by the technology. Therefore, they discontinued before completing any full VR sessions. The remaining 10 participants (91%) completed the 4-week intervention and follow-up assessments. All completers achieved the target of eight VR sessions (100% adherence), and no further dropouts or protocol deviations were observed. Approximately half of the participants required minor technical support (such as help with VR content connection or how to use Meta Quest 2) during the first week, which was resolved via e-mail, an online Question & Answer manual for this research, or a brief phone call. None of the patients withdrew due to adverse effects.

### Baseline Characteristics

Table 1 presents the baseline demographic and clinical characteristics of the 10 participants. The mean age was 35 years (SD 8.8, range 21–49), with 60% females and 40% males. All the participants were of Japanese ethnicity. The primary diagnosis categories were anxiety/adjustment in 4 participants (40%), mood disorders in 3 (30%), somatic symptom disorder in 2 (20%), and insomnia disorder in 1 (10%). “Anxiety/adjustment” included generalized anxiety disorder, adjustment disorder, and social anxiety; “mood” included dysthymia and bipolar II. All patients were in active treatment for their conditions; eight (80%) were taking at least one medication (typically a sedative-hypnotic or anxiolytic) for insomnia or anxiety, whereas two (20%) were not on any psychoactive medication. None of the participants had prior experience with VR therapy or formal behavioral therapy for insomnia. Baseline symptom scores indicated moderate severity overall: the group mean GAD-7 was 9.9 (SD 3.2), PHQ-9 was 12.6 (SD 3.4), and PSQI was 10.3 (SD 1.6), reflecting significant insomnia (all participants scored ≥6 on PSQI) and mild-to-moderate anxiety or depressive symptoms. Individually, 50% of participants fell in the “moderate anxiety” range (GAD-7 scores 10–14), 40% in the mild range (5–9), and 10% met the severe threshold (≥15; score = 15). All 10 had PSQI ≥8 (well above the clinical cutoff of>5 for insomnia), consistent with moderate to severe sleep disturbance.

**Table 1.** Participant baseline characteristics (N=10)

| Characteristic | Value |
| --- | --- |
| Age, years | 35.0±8.8 (mean±standard deviation) |
| Female sex, n (%) | 6 (60%) |
| Ethnicity, n (%) | Japanese, 10 (100%) |
| Primary diagnosis category, n (%) | Anxiety/adjustment 4 (40%); Mood 3 (30%); Somatic 2 (20%); Insomnia 1 (10%) |
| On psychiatric medication, n (%) | 8 (80%) |
| GAD-7 anxiety score (0–21) | 9.9±3.2 (moderate) |
| PHQ-9 depression score (0–27) | 12.6±3.4 (moderate) |
| PSQI global sleep quality score (0–21) | 10.3±1.6 (poor) |

Abbreviations: GAD-7, Generalized Anxiety Disorder-7; PHQ-9, Patient Health Questionnaire-9;
PSQI, Pittsburgh Sleep Quality Index; n, number of participants.

### Acceptability and Safety

The participants considered the home-based VR relaxation intervention acceptable and easy to incorporate into their routines. All 10 completers finished the prescribed eight sessions within 4 weeks, and half of the sample expressed interest in continuing VR use beyond the study. In the post-intervention VR experience survey, the overall satisfaction and immersion were high.

Table 2 presents the distribution of responses to the five user-experience questions, and Fig 4 shows the item-level responses. Most participants rated the VR system as easy to use and the content as engaging. For example, 40% rated “mostly satisfied” and 10% “very satisfied” with the ease of VR setup, and 30% selected a mid-score (3/5); no one rated the ease as “very difficult”, and 20% selected 2/5 (indicating slight difficulty with setup). For immersion (“being present”), 90% rated ≥3/5 (pre-specified as moderate-to-high), and 10% rated 2/5 (below moderate). The quality of the scenic content was similarly well-received, with 60% giving a 3/5 (“moderately satisfied”), 20% giving 4/5 (“mostly satisfied”), and 10% giving 5/5 (“very satisfied”). Minor discomfort occurred in 3/10 participants (30%; transient eyestrain or dizziness). No serious adverse events occurred. No sessions were aborted due to simulator sickness, and no participant reported any persistent increase in anxiety. The single dropout cited feeling stressed by the VR equipment, underscoring the fact that a small subset of participants (perhaps those who are less tech-comfortable) may struggle with VR. However, among the completers, no further dropouts were observed owing to adverse effects, and the overall safety was very favorable. Aside from the one participant who withdrew due to feeling overwhelmed by the technology (a non-physical issue), no adverse events led to discontinuation.

**Fig 4.**
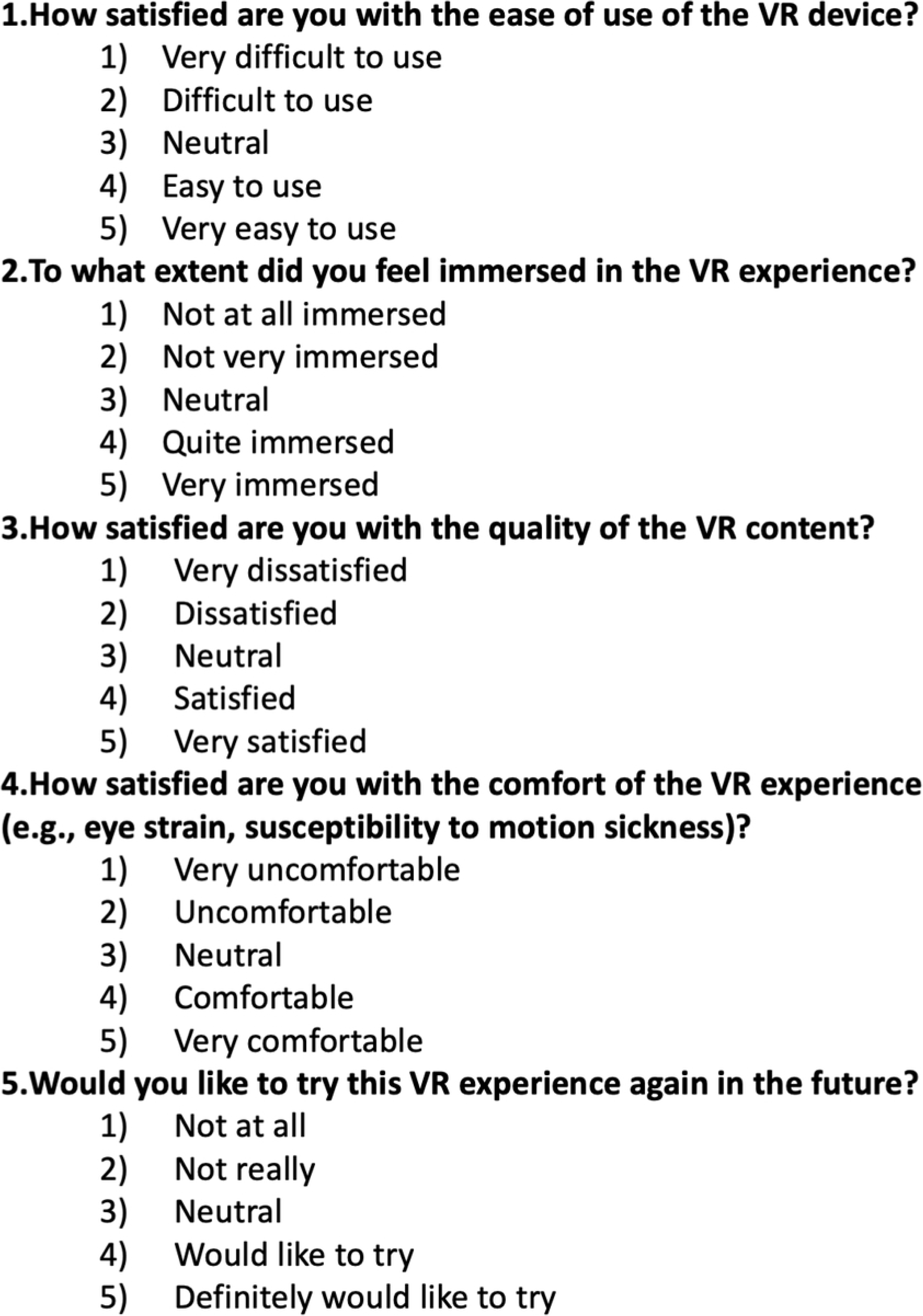
Virtual reality experience survey (post-intervention). Five items were administered after the 4-week intervention using a 5-point Likert scale (1=worst/strongly negative, 5=best/strongly positive).

**Table 2.**
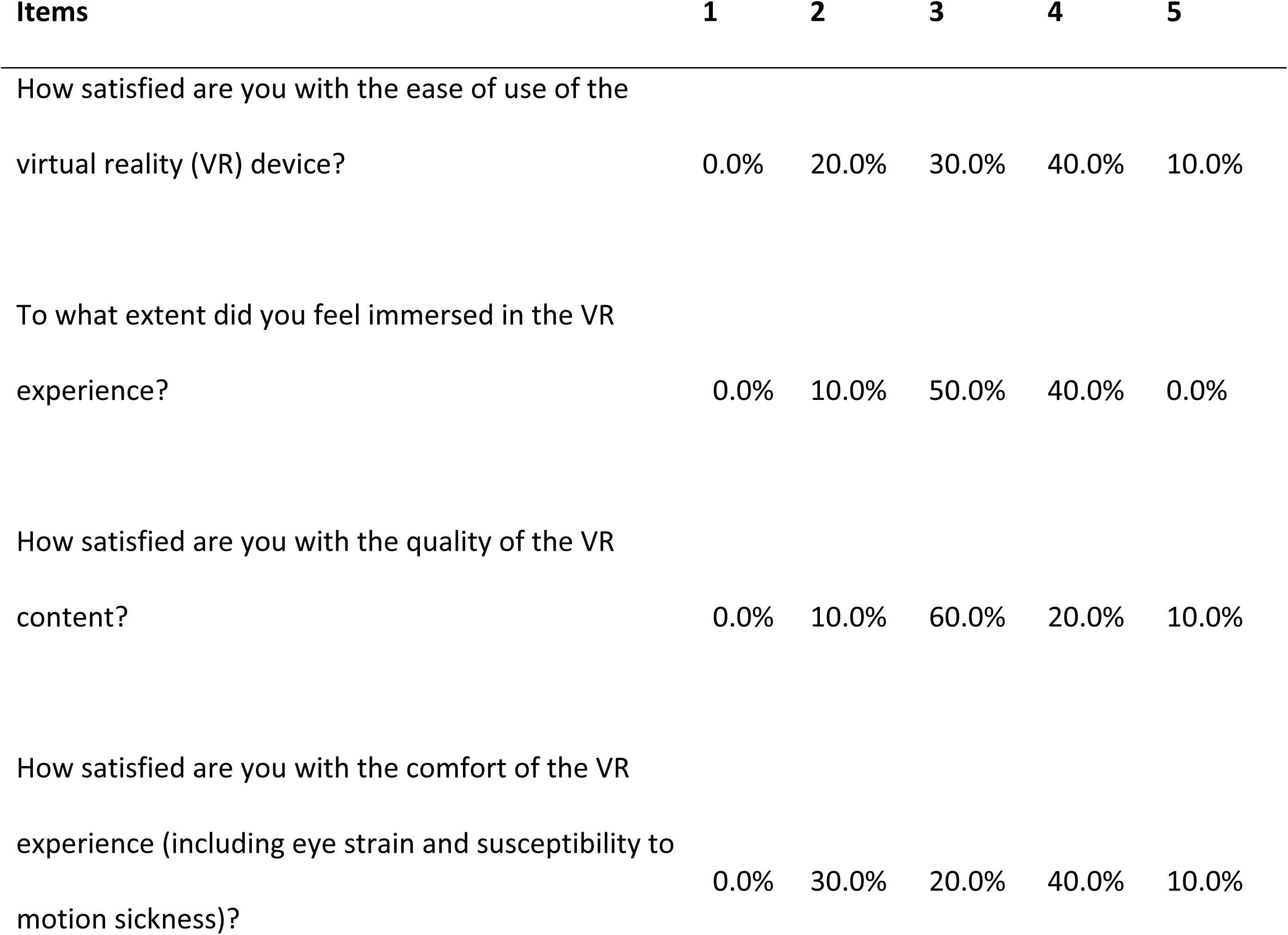
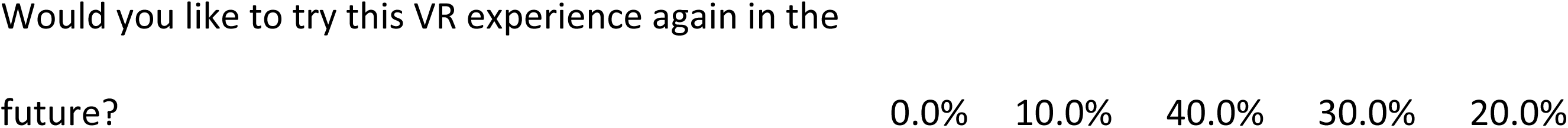
Distribution of virtual reality experience survey responses (%, N=10). Columns show the percentage selecting each Likert option (1–5), by item. Higher scores indicate more favorable ratings. Totals may not equal 100% due to rounding.

### Clinical Outcomes (Anxiety, Depression, and Sleep)

All available pre- and post-intervention symptom scores for the 10 participants are shown in Fig 5. Each outcome improved in the expected direction after the 4-week VR intervention. The mean group scores and statistical comparisons are presented in Table 3.

**Fig 5.**
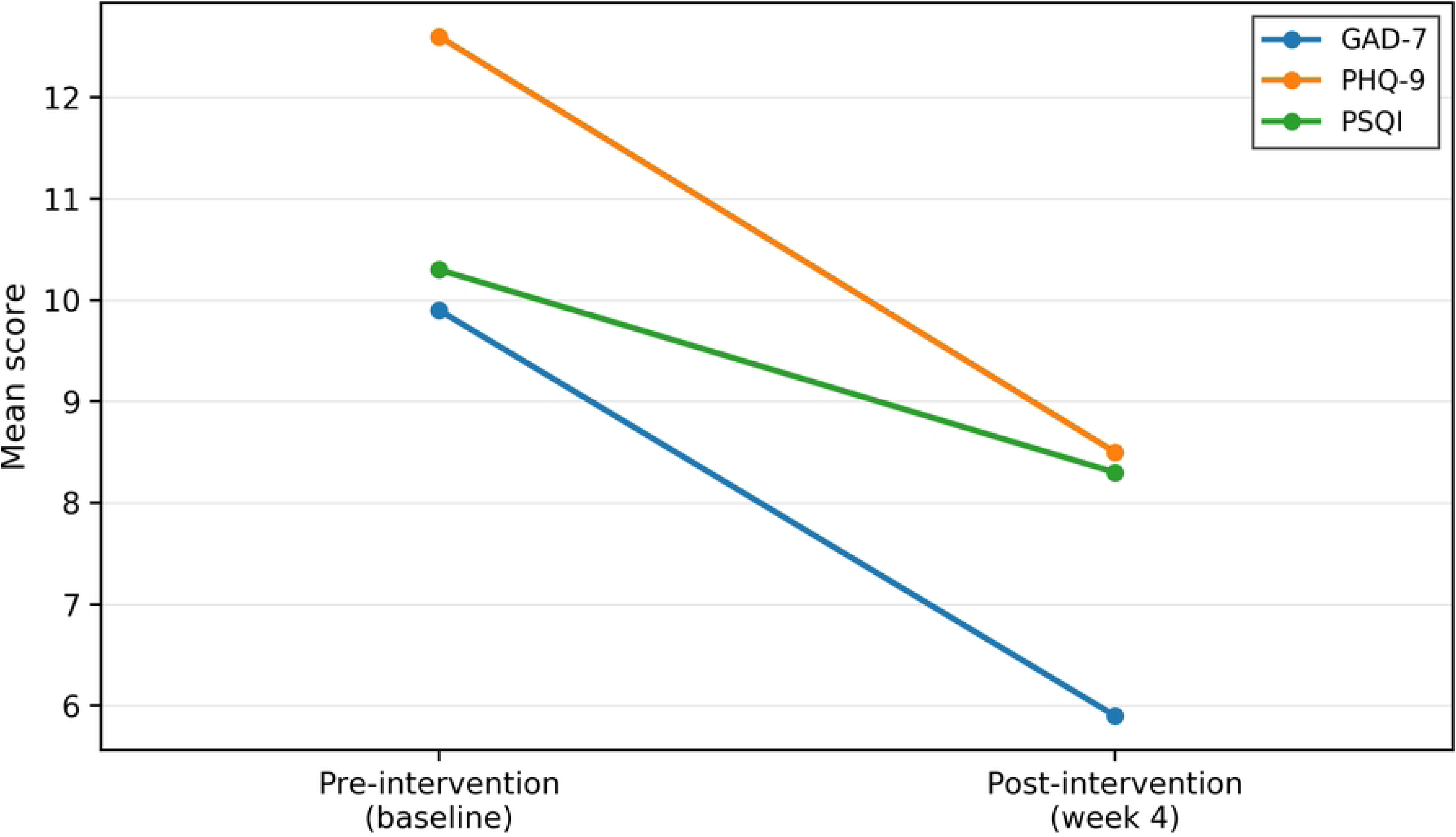
Group mean GAD-7, PHQ-9, and PSQI at pre- and post-intervention. Lines connect group means at pre- and post-intervention (week 4). Higher scores indicate worse symptoms. Completers: n = 10. Abbreviations: GAD-7, Generalized Anxiety Disorder-7; PHQ-9, Patient Health Questionnaire-9; PSQI, Pittsburgh Sleep Quality Index;

**Table 3.** Pre- and post-intervention symptom scores (mean±standard deviation) and mean changes (N=10).

| Outcome | Baseline<br>(Pre) | 4-week<br>(Post) | Mean change (Post–Pre),<br>95% confidence interval | Cohen’s dz, 95%<br>confidence<br>interval |
| --- | --- | --- | --- | --- |
| GAD-7 anxiety<br>score (0–21) | 9.9±3.2 | 5.9±1.4 | –4.0, –6.00 to –2.00 | 1.43, 0.96 to 2.64 |
| PHQ-9 depression<br>score (0–27) | 12.6±3.4 | 8.5±3.0 | –4.1, –7.32 to –0.88 | 0.91, 0.41 to 1.86 |
| PSQI sleep quality<br>global score (0–21) | 10.3±1.6 | 8.3±2.3 | –2.0, –3.47 to –0.53 | 0.95, 0.35 to 1.72 |

Abbreviations: GAD-7, Generalized Anxiety Disorder-7; PHQ-9, Patient Health Questionnaire-9; PSQI, Pittsburgh Sleep Quality Index; CI, confidence interval; dz, within-subject Cohen’s d; n, number of participants.

On the GAD-7 anxiety scale, scores decreased in nine out of 10 individuals. Group mean GAD-7 dropped from 9.9 at baseline to 5.9 at week 4, an average reduction of –4.0 points (95% CI – 6.00 to –2.00). This corresponds to an approximately 40% improvement in anxiety severity, and a large within-subject effect size was Cohen’s dz=1.43 (95% CI 0.96–2.64) indicating a reduction in anxiety. Fig 6 shows that most participants’ GAD-7 trajectories were downward-sloping.

**Fig 6.**
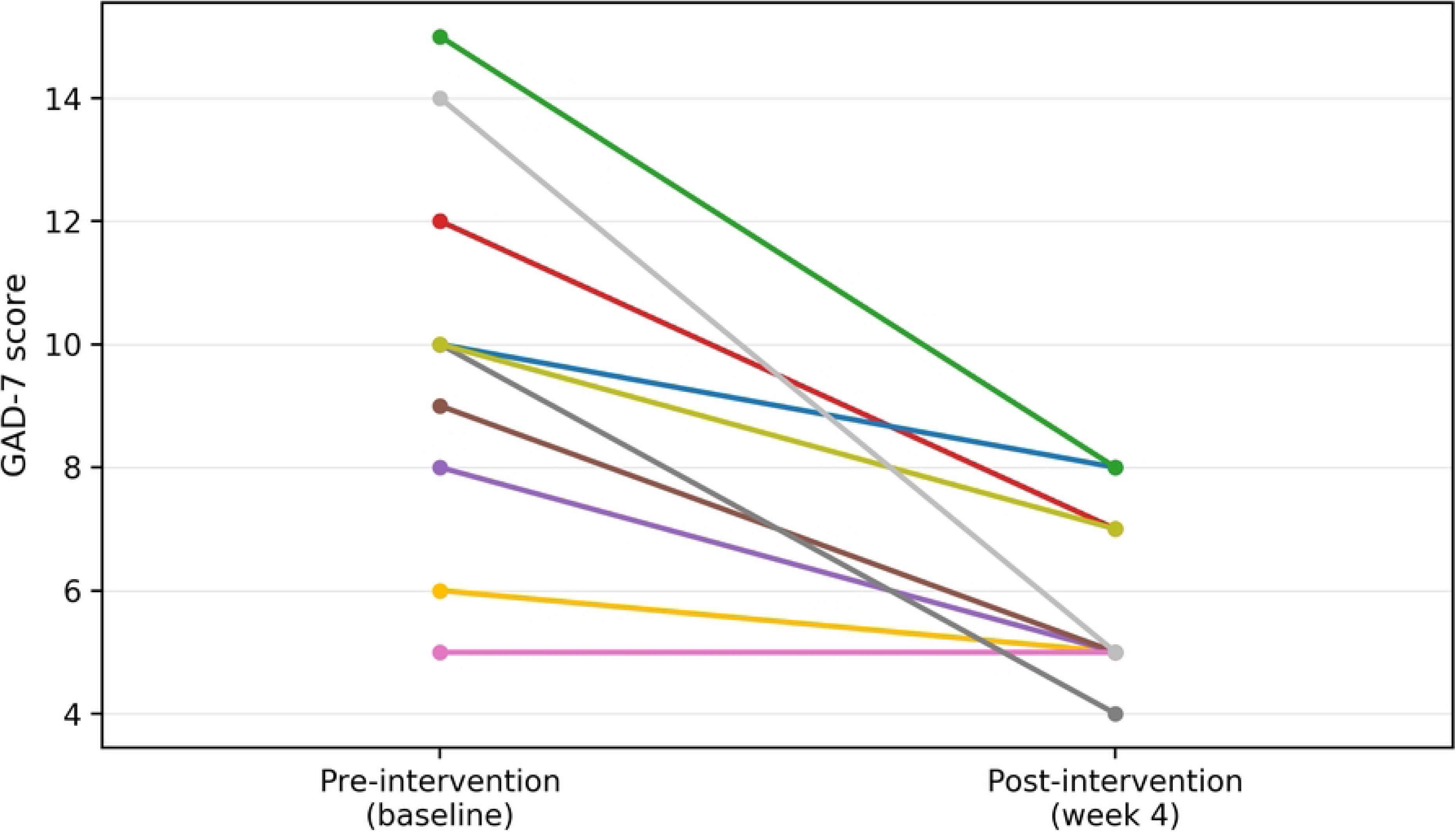
Individual pre–post trajectories for GAD-7. Each line shows one participant (n = 10). No overlapping pre–post pairs. Abbreviations: GAD-7, Generalized Anxiety Disorder-7.

Overall, 7/10 participants (70%) achieved a clinically meaningful anxiety reduction of ≥3 points, and by study end none of the participants remained in the moderate or severe anxiety range (GAD-7≥10). In fact, 90% had shifted into the “mild” anxiety category (scores 5–9), and one patient’s score fell to 4, indicating minimal anxiety. This pattern suggests that VR relaxation may have helped reduce the participants’ excessive worry and arousal.

Depressive symptoms (PHQ-9) also improved substantially. The mean PHQ-9 score decreased from 12.6 baseline to 8.5 week 4, for an average change of –4.1 points (95% CI –7.3 to –0.9). The within-subject effect size for depression improvement was Cohen’s dz=0.91 (95% CI 0.41– 1.86). As Fig 7 indicates, individual depression scores showed more variability than anxiety scores. However, seven of 10 participants had lower PHQ-9 scores after the intervention. Four of 10 participants achieved a ≥5-point drop in PHQ-9. The proportion of patients in the moderate-to-severe depression range (PHQ-9 ≥10) fell from 70% at baseline to 50% at follow-up. Notably, one participant (10%) had minimal depressive symptoms (PHQ-9<5) after the intervention, whereas none had scores that were low at baseline. This suggests that for a subset of patients, VR-based relaxation was associated with marked relief of mood symptoms. One participant had essentially no change in PHQ-9 (remaining at 8 points). Two participants worsened slightly (increases of 1 and 2 points), and no participant had a 5-point or greater increase, indicating no clinically significant worsening by that threshold.

**Fig 7.**
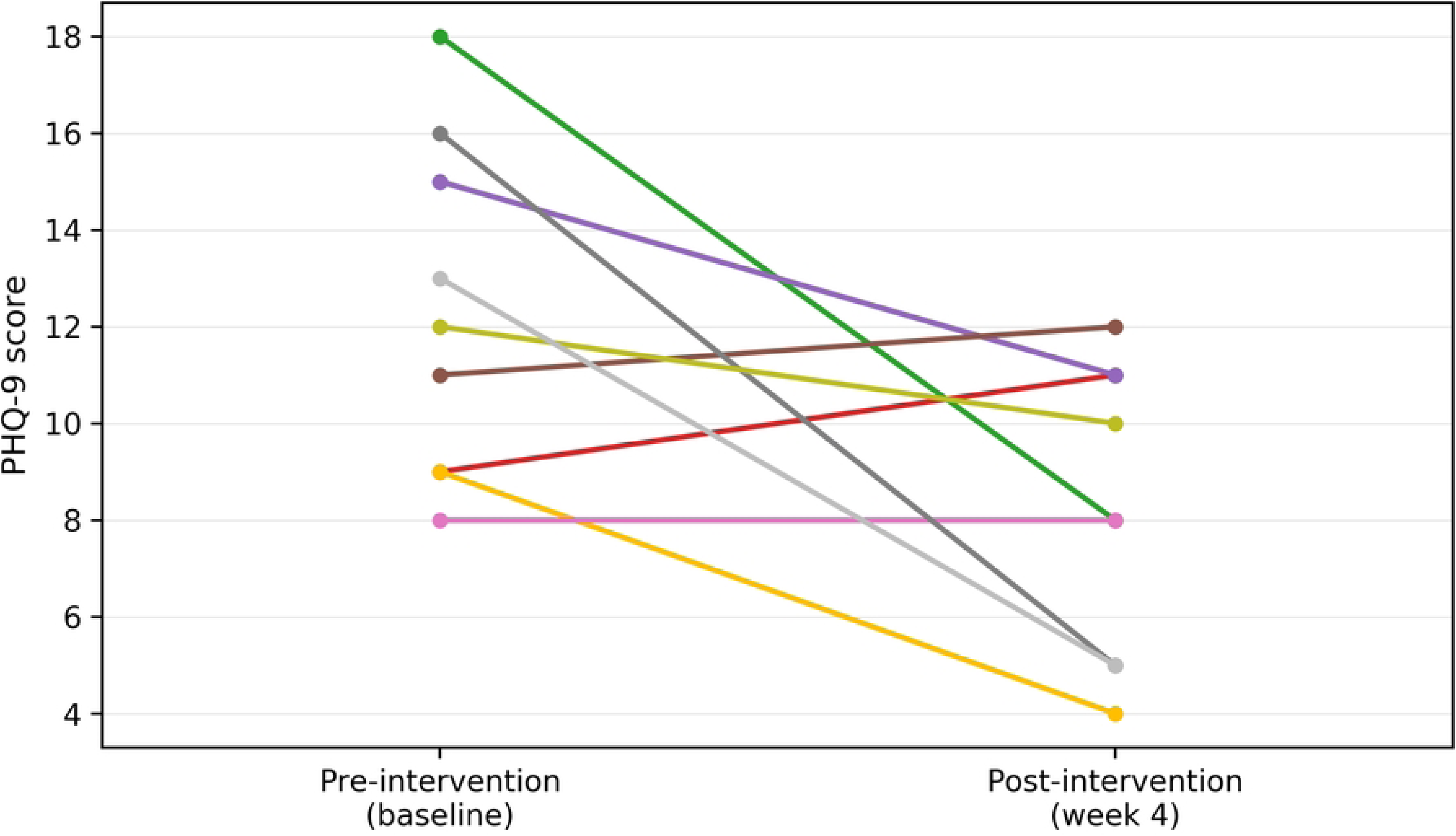
Individual pre–post trajectories for PHQ-9. Ten participants are plotted; two participants share identical pre–post values, so 9 lines are visible. Abbreviations: PHQ-9, Patient Health Questionnaire-9.

Sleep quality (measured by the PSQI) showed a more modest improvement. Mean global PSQI decreased from 10.3 at baseline to 8.3 at post-intervention (week 4; mean difference −2.0, 95% CI −3.5 to −0.5; Fig 8). Eight of ten participants improved; two worsened by 1–2 points. The within-participant effect size (Cohen’s *dz*) was 0.95. At week 4, all PSQI scores remained >5 (good-sleep cutoff ≤5); two participants reached 6. Reductions were largest for subjective sleep quality (C1) and sleep latency (C2), with smaller changes for sleep disturbances (C5) and daytime dysfunction (C7) (see S1 Table). One participant with a +2-point PSQI increase had a −4-point GAD-7 reduction, suggesting sleep variability unrelated to anxiety change.

**Fig 8.**
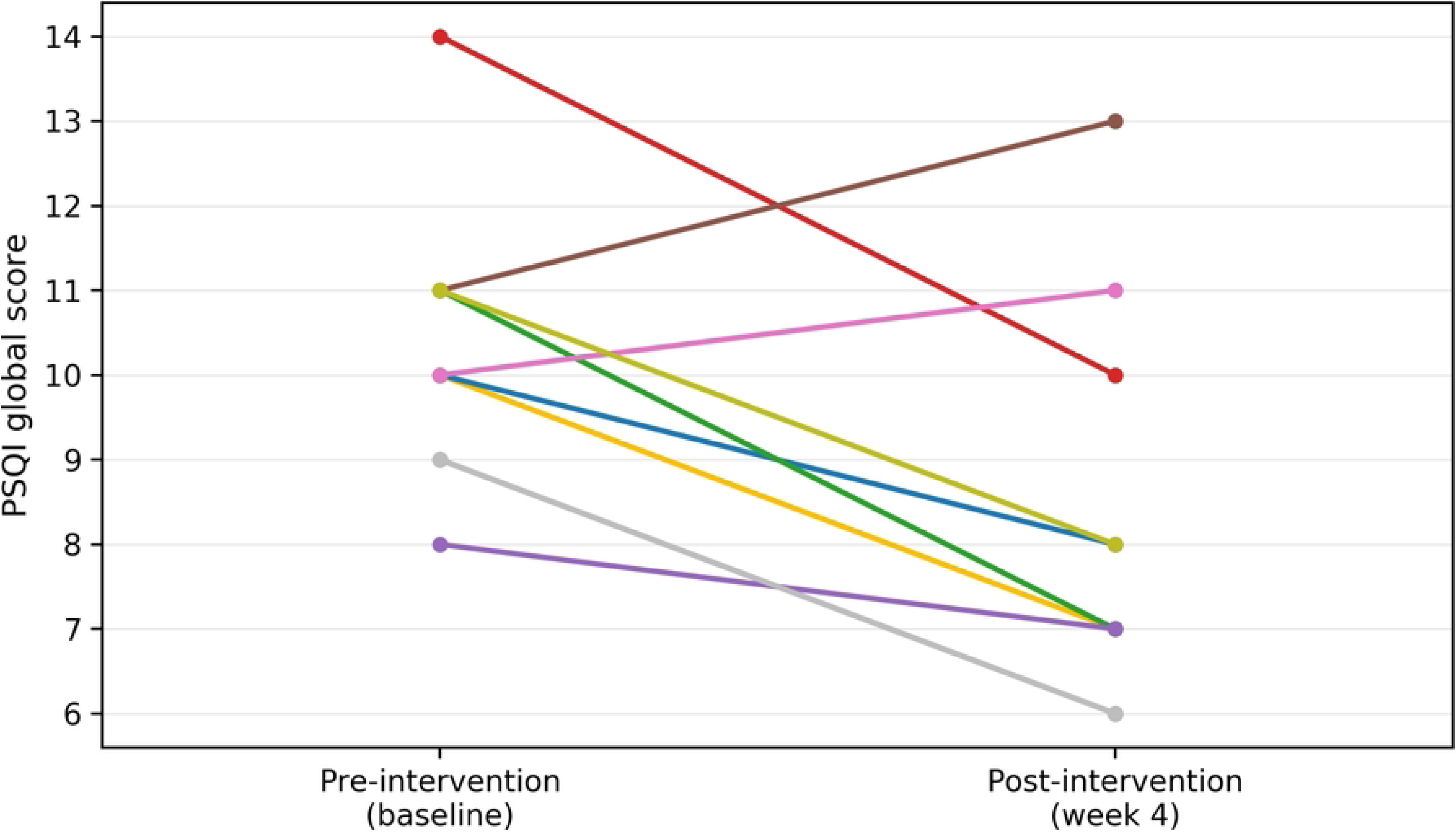
Individual pre–post trajectories for PSQI. Ten participants are plotted; two participants share identical pre–post values, so 9 lines are visible. Higher PSQI indicates worse sleep. Abbreviations: PSQI, Pittsburgh Sleep Quality Index.

In summary, the VR scenic relaxation intervention was associated with clinically meaningful reductions in anxiety and depression, and a trend toward better sleep quality. No formal hypothesis testing was performed (given the pilot nature). However, the 95% CIs for mean changes in the GAD-7 and PHQ-9 scores did not cross zero, and even the sleep score CI barely excluded no change. These improvements, observed without any new pharmacotherapy or intensive counseling, suggest that a brief course of VR-guided relaxation may provide measurable relief from symptoms in participants with chronic insomnia and comorbid anxiety. We emphasize that these clinical results are exploratory; the study was not powered by efficacy, but the effect sizes were moderate to large. For instance, a mean GAD-7 score drop of 4 points is consistent with a clinically meaningful change and suggests its potential utility as an adjunct to usual care. Similarly, depression and sleep improvements were more variable, which aligned with improvements observed in other mind–body interventions for this population. Together with high acceptability and adherence, these outcome trends support further research to formally test VR relaxation as a therapeutic option for insomnia and anxiety. The next step will be a controlled trial to determine if these pre–post improvements can be definitively attributed to the VR intervention.

## Discussion

### Key Findings

This exploratory pilot study demonstrated that a 4-week self-administered VR scenic relaxation intervention was feasible, safe, and acceptable for participants with chronic insomnia and co-occurring anxiety. We achieved a high recruitment and completion rate; 91% of the enrolled participants completed all sessions and assessments, and adherence to the 8-session protocol was 100% among completers. These feasibility results are comparable to other recent VR initiatives in mental health; for example, a 4-week, group-based VR gaming program for adolescents with physical disabilities reported approximately 67% retention [12]. In our adult outpatient sample, motivation to engage in VR therapy was high, and only one person (9%) withdrew early because of technological discomfort. No serious AEs were observed. Minor headset-related discomfort (eye strain, susceptibility to motion sickness) was reported by 30% (VR experience survey Q4). This safety and acceptability profile aligns with prior VR relaxation studies in outpatient settings and home-delivered interventions [16,17,25], and is consistent with other home or self-guided VR approaches [13,16]. Participants reported the VR experience as acceptable and engaging. Willingness to reuse was moderate: 5/10 (50%) selected 4–5/5, 4/10 (40%) selected 3/5, and 1/10 (10%) selected 2/5. These ratings indicate interest in continued use for some, but not all, participants. In this small, uncontrolled pilot, home-based VR scenic relaxation appeared practical and well received in this sample.

Regarding clinical outcomes, our uncontrolled pre–post analysis indicated improvements in anxiety, depression, and sleep after the VR intervention. Anxiety symptoms (GAD-7) decreased by approximately 4 points on average, which meets the published minimal clinically important difference (approximately 4 points) [26] and is consistent with a potential one-category shift on standard cut points — moderate (10–14) to mild (5–9) [14]. Depression scores (PHQ-9) also decreased (approximately 4-point improvement), and self-reported sleep disturbances (PSQI) improved modestly (approximately 2-point mean decrease) over 4 weeks. Notably, seven out of 10 participants showed improvement across the anxiety, depression, and sleep domains, suggesting a potential broad benefit, albeit in this small sample. The within-subject effect sizes were in the moderate-to-large range (Cohen’s dz=0.9–1.4 for the three measures), further underscoring the potential efficacy of the VR intervention despite the small sample. While one must be cautious in interpreting uncontrolled data, the consistency of improvements, particularly the robust reduction in anxiety [27], supports our hypothesis that immersive VR relaxation may help reduce pre-sleep arousal and related symptoms in this population. This pattern aligns with studies showing that immersive VR (including meditation and biofeedback) improves sleep-related outcomes and autonomic function [28,29]. It is also consistent with the theoretical rationale for bedtime VR to reduce pre-sleep hyperarousal [30].

### Implications and Comparison to Prior Research

These preliminary results have several implications regarding the role of VR in mental health and sleep therapy. First, it extends prior VR research into a new context, showing that a self-administered home-based VR relaxation intervention is workable and beneficial for participants with clinically significant insomnia and anxiety. Many early VR interventions require in-clinic sessions or therapist guidance, whereas our approach is largely independent and remote. This at-home model is potentially scalable and cost-effective, reducing barriers to non-pharmacological care for insomnia and anxiety (time, mobility, stigma) by allowing patients to complete sessions on their own schedule after basic training. The high user satisfaction we observed suggests that the participants observed value in VR as a self-care tool. Immersive VR may serve as a “gateway” for relaxation or mindfulness practices among those who otherwise struggle with traditional techniques. The engaging nature of VR makes it easier to achieve a calm, meditative state compared with audio-only meditation – an insight echoed by our participants and also observed in home or remote VR relaxation interventions [7,13,16].

Another important implication is the transdiagnostic potential of VR relaxation. Diagnoses in the cohort were heterogeneous: 40% anxiety or adjustment, 30% mood, 20% somatic symptom disorder, and 10% insomnia disorder. Most participants showed reductions in anxiety and improved sleep. This suggests that VR-guided relaxation targets common underlying mechanisms, such as hyperarousal, rumination, and difficulty unwinding, which cut across diagnostic categories. In other words, VR relaxation can provide a general calming effect that is broadly applicable rather than disorder-specific. This aligns with the notion that many psychiatric conditions share an elevated arousal component (whether cognitive or physiological), which interventions such as VR can help downregulate. Clinically, VR relaxation could be prescribed as an adjunctive therapy for anxiety reduction and insomnia improvement alongside standard treatments in diverse patient groups. In our study, 80% of the participants were already on medication. However, VR was a helpful complement, empowering them to actively manage their nighttime anxiety and insomnia in a self-directed manner. Notably, none of our participants experienced deterioration in their symptoms while using VR, and even those who had minimal improvement did not experience any harm. Safety and non-interference with usual care are critical for providers and patient confidence in adopting VR. Retention was 91%; all completers finished 8/8 sessions, and no SAEs occurred. On this basis, home-based VR relaxation is safe and feasible as an adjunct to usual care, in line with the feasibility reported in other home or telehealth VR interventions that also report benefits for sleep and mood [13,16].

Our findings also contribute to the discussion on whether the observed improvements were truly due to the VR intervention versus nonspecific effects or natural recovery. Without a control group, we could not definitively separate the impact of VR from the placebo response or the passage of time. However, the magnitude and pattern of the results support the possibility that the VR intervention provided a therapeutic benefit. The approximately 40% reduction in GAD-7 scores within 4 weeks is larger than what we would expect from usual care or spontaneous improvement alone over such a short period, particularly considering that participants were stable on their routine treatments when entering the study. Participants reported moderate-to-high satisfaction and immersion, and exploratory pre–post analyses showed mean improvements in anxiety, depression, and sleep scores. This subjective impression is supported by emerging evidence from controlled trials. For example, a randomized study on chronic insomnia reported greater improvements in PSQI and anxiety when VR therapy was added to standard care than with standard care alone [7]. Additional randomized and sham-controlled studies in related contexts showed that immersive VR outperforms guided imagery or audio-only controls on state anxiety, mood, and sleep-related outcomes [28,31]. Similarly, a retrospective clinical study of 103 patients with anxiety and insomnia reported immediate anxiety relief and sleep improvement after brief VR-integrated relaxation sessions [8]. These convergent findings from prior research reinforce the likelihood that our observed improvements were not merely a regression to the mean; VR had a tangible therapeutic impact by immersing patients in a calming virtual environment and evoking a relaxation response. Nonetheless, we acknowledge that some improvements could reflect expectancy or the general therapeutic milieu of participating in a study. We attempted to minimize this by keeping other aspects of care constant and making the VR intervention self-guided (with minimal clinician contact). However, only a controlled trial could fully address this question.

### Limitations

This study had some limitations inherent to a small, single-arm pilot study. Design and interpretability: The study had 10 completers and no control group, making the effect estimates imprecise and unable to be causally attributed to the VR intervention. Natural symptom fluctuations, regression to the mean, and expectancy or placebo effects may have contributed to these findings. Setting and generalizability: The participants were recruited from a single specialty clinic, and the cohort was relatively young (predominantly 20–40 years old), which limited the external validity of the study. Acceptance, usability, and effects may differ in older adults or populations less familiar with head-mounted displays, highlighting the need for ongoing trials in these populations [11]. Follow-up window: Outcomes were assessed immediately after a 4-week intervention; therefore, durability was unknown. Longer follow-up is required to determine whether gains persist without continued VR use or whether periodic “booster” sessions are needed to maintain improvements. Concomitant treatments: Eight of the ten participants were taking psychotropic medication at baseline and continued on stable regimens during the study. Because pharmacotherapy was maintained rather than randomized or withdrawn, the effects of medication on pre–post changes cannot be excluded. Outcome measurement: We relied on self-report questionnaires for sleep and anxiety, lacking objective sleep indices (such as actigraphy or polysomnography) or physiological arousal measures (such as heart rate or heart rate variability and cortisol), which limits the ability to validate subjective reports. As a result, candidate mechanisms, such as reductions in nocturnal hyperarousal, remain speculative; in short trials, subjective gains can precede changes in objective sleep, emphasizing the need to include objective endpoints in future studies [28]. Intervention content and personalization: VR content was restricted to two 360° natural scenes delivered in a fixed sequence, which may have limited user engagement and the potential effectiveness of the intervention. This simplicity favored usability but may not maximize engagement across users; we did not compare content types (such as interactive or guided-meditation VR) or allow personalization (including scene choice and biofeedback), such that we could not assess whether tailoring would enhance outcomes. Precision and scope: The small sample size limited statistical precision (wide CIs) and precluded moderator or subgroup analyses (including baseline severity or medication use). Collectively, these constraints mean that the findings should be viewed as preliminary and hypothesis-generating, and confirmation in larger, controlled trials with longer follow-up periods, objective and mechanistic endpoints, and comparative VR content is warranted.

### Future Directions

The pilot safety and adherence support a sample-size planning RCT to estimate the treatment effects and variability under active control conditions. A controlled trial could compare the VR relaxation intervention to an active control condition, for example, guided imagery [31] or a sham-VR model [32], to account for nonspecific effects. A sham-controlled randomized trial will show how much of the benefit comes from immersion rather than from nonspecific effects. It would also be informative to compare VR relaxation with gold standard treatments, such as cognitive-behavioral therapy for insomnia (CBT-I), or to test VR as an adjunct that augments the effects of established therapies. Compare CBT-I plus home VR with CBT-I alone in a randomized trial and quantify the incremental improvement. Prior VR trials rechecked outcomes approximately 3–6 months after treatment [32,33] and used brief refreshing or booster contact [34]. Following this precedent, we plan to include 3- and 6-month follow-ups (GAD-7, PHQ-9, and PSQI) and test a monthly booster schedule. It may also be useful to track how participants incorporate VR into their routine once the study ends. Many intended to continue using it; therefore, a follow-up could determine if and how it is performed. Another avenue is to test VR in more severe or refractory cases; for example, patients who cannot tolerate medications or those with chronic insomnia who do not fully respond to CBT-I might particularly benefit from an immersive relaxation tool. Additionally, research on the optimal level of human support required for VR interventions is warranted. Our pilot provided technical support but was otherwise self-guided. Future work could explore whether adding minimal clinician coaching or remote monitoring improves adherence or outcomes. Identifying the most scalable yet effective way to deliver VR (fully self-guided vs. some support) will be the key to its implementation. A future cost-effectiveness analysis is also warranted. We intend to estimate the incremental cost-effectiveness (from a payer perspective) by accounting for device, logistics, and personnel costs against quality-adjusted life year gains, once efficacy is established in a larger trial. As VR hardware becomes more affordable, one can envision clinics maintaining a set of headsets to loan to patients (similar to the model of our study) or patients purchasing their own if the therapeutic value justifies it. Analyses of whether VR relaxation can reduce healthcare utilization (such as decrease medication needs or clinic visits) and improve functional outcomes will help establish its value proposition in the digital mental health toolkit, with home or telehealth VR interventions providing encouraging precedents for sustained adherence and mood or sleep benefits [7,16,25]. In summary, future research should focus on confirming its efficacy in a controlled setting, optimizing the intervention (content and delivery), and understanding for whom and under what circumstances VR relaxation works best. If the effects are reproduced under active control, VR relaxation would be a viable adjunct for improving sleep and mood outcomes in outpatient care.

## Conclusion

This exploratory feasibility pilot considered a 4-week home-based VR scenic relaxation intervention feasible, safe, and acceptable for outpatients with chronic insomnia and anxiety symptoms (completion 91%; adherence 100%). No SAEs occurred, and only mild transient discomfort was reported. Exploratory outcomes suggested improvements in anxiety, depression, and sleep. Given the single-arm design, these findings are preliminary. However, they support conducting a larger, controlled trial to confirm efficacy and refine implementation.

## Data Availability

All de-identified participant-level data underlying the results presented in this study are available in the Zenodo repository (DOI: 10.5281/zenodo.17338176). The deposit includes: (i) pre-/post-intervention outcomes (prepost_dataset.csv) (ii) VR experience survey responses (vr_survey_dataset.csv) (iii) baseline characteristics used for Table 1 (background_dataset.csv) and (iv) a data dictionary (S1_Data_Dictionary_3datasets.xlsx) describing all variables and coding. Figure source files/underlying values for the summary graphics are also provided. To protect privacy, all direct identifiers and exact dates have been removed. There are no access restrictions.

https://zenodo.org/records/17338176

## Acknowledgments

We thank Ms. Hayashi of the Clinical Research and Trial Center, Juntendo University, Tokyo, Japan, for expert coordination and operational support in conducting this intervention, including participant screening, data input, and management.

## Data Availability

All de-identified data underlying the results (pre/post outcomes and VR experience survey) and figure source files are available at Zenodo (doi: 10.5281/zenodo.17338176).

## Supporting information

S1 Checklist. TREND reporting checklist.

S1 Protocol (Japanese, original). CRB-approved clinical trial protocol (Version 2.0, approval J24-007).

S2 Protocol (English translation). CRB-approved clinical trial protocol (Version 2.0, approval J24-007).

S1 Table. PSQI component scores (C1–C7) at baseline and 4 weeks, and mean change (N=10).

## References

1. Itani O, Kaneita Y, Tokunaga H, Jike M, Murata A, et al. Nationwide epidemiological study of insomnia in Japan. Sleep Med. 2016;25:130–138. doi:10.1016/j.sleep.2016.05.013.

2. Oka Y, Murasaki Y, Mori S. Current trends in sleep hygiene promotion and insomnia treatment: Cooperation among medical professionals and collaboration with pharmaceutical industry. J Sleep Environ. 2025;19: 50–59. doi: 10.60259/jsleepenvi.19.1_50.

3. Riemann D, Espie CA, Altena E, Arnardottir ES, Baglioni C, Bassetti CLA, et al. The European Insomnia Guideline: An update on the diagnosis and treatment of insomnia 2023. J Sleep Res. 2023;32(6):e14035. doi: 10.1111/jsr.14035.

4. Carl E, Stein AT, Levihn-Coon A, Pogue JR, Rothbaum B, Emmelkamp P, et al. Virtual reality exposure therapy for anxiety and related disorders: A meta-analysis of randomized controlled trials. J Anxiety Disord. 2019;61: 27–36. doi: 10.1016/j.janxdis.2018.08.003.

5. Riches S, Jeyarajaguru P, Taylor L, Fialho C, Little J, Ahmed L, et al. Virtual reality relaxation for people with mental health conditions: A systematic review. Soc Psychiatry Psychiatr Epidemiol. 2023;58: 989–1007. doi: 10.1007/s00127-022-02417-5.

6. Meshkat S, Edalatkhah M, Di Luciano C, Martin J, Kaur G, Hee Lee G, et al. Virtual reality and stress management: A systematic review. Cureus. 2024;16: e64573. doi: 10.7759/cureus.64573.

7. Wan Y, Gao H, Zhou K, Zhang X, Xue R, Zhang N. Virtual reality improves sleep quality and associated symptoms in patients with chronic insomnia. Sleep Med. 2024;122: 230–236. doi: 10.1016/j.sleep.2024.08.027.

8. Zhou H, Chen C, Liu J, Fan C. Acute augmented effect of virtual reality (VR)-integrated relaxation and mindfulness exercising on anxiety and insomnia symptoms: A retrospective analysis of 103 anxiety disorder patients with prominent insomnia. Brain Behav. 2024;14: e70060. doi: 10.1002/brb3.70060.

9. de Zambotti M, Yuksel D, Kiss O, Barresi G, Arra N, Volpe L, et al. A virtual reality-based mind–body approach to downregulate psychophysiological arousal in adolescent insomnia. Digit Health. 2022;8: 20552076221107887. doi: 10.1177/20552076221107887.

10. Ma J, Li H, Liang C, Li S, Liu Z, Qu C. A brief virtual reality-based mindfulness intervention can improve olfactory perception while reducing depression and anxiety symptoms in university students. Humanit Soc Sci Commun. 2025;12: 294. doi: 10.1057/s41599-025-04584-7.

11. Cinalioglu K, Lavín P, Bein M, Lesage M, Gruber J, Se J, et al. Effects of virtual reality guided meditation in older adults: The protocol of a pilot randomized controlled trial. Front Psychol. 2023;14: 1083219. doi: 10.3389/fpsyg.2023.1083219.

12. Lai B, Young R, Craig M, Chaviano K, Swanson-Kimani E, Wozow C, et al. Improving social isolation and loneliness among adolescents with physical disabilities through group-based virtual reality gaming: Feasibility pre–post trial study. JMIR Form Res. 2023;7: e47630. doi: 10.2196/47630.

13. Pallavicini F, Orena E, di Santo S, Greci L, Caragnano C, Ranieri P, et al. A virtual reality home-based training for the management of stress and anxiety among healthcare workers during the COVID-19 pandemic: Study protocol for a randomized controlled trial. Trials. 2022;23: 451. doi: 10.1186/s13063-022-06337-2.

14. Spitzer RL, Kroenke K, Williams JBW, Löwe B. A brief measure for assessing generalized anxiety disorder: The GAD-7. Arch Intern Med. 2006;166: 1092–1097. doi: 10.1001/archinte.166.10.1092.

15. Buysse DJ, Reynolds CF III, Monk TH, Berman SR, Kupfer DJ. The Pittsburgh Sleep Quality Index: A new instrument for psychiatric practice and research. Psychiatry Res. 1989;28: 193–213. doi: 10.1016/0165-1781(89)90047-4.

16. Riches S, Kaleva I, Nicholson SL, Payne-Gill J, Steer N, Azevedo L, et al. Virtual reality relaxation for stress in young adults: A remotely delivered pilot study in participants’ homes. J Technol Behav Sci. 2024;9: 771–783. doi: 10.1007/s41347-024-00394-x.

17. Chang E, Kim HT, Yoo B. Virtual reality sickness: A review of causes and measurements. Int J Hum Comput Interact. 2020;36: 1658–1682. doi: 10.1080/10447318.2020.1778351.

18. Kroenke K, Spitzer RL, Williams JBW. The PHQ-9: Validity of a brief depression severity measure. J Gen Intern Med. 2001;16: 606–613. doi: 10.1046/j.1525-1497.2001.016009606.x.

19. International Conference on Harmonisation (ICH). Clinical safety data management: definitions and standards for expedited reporting (E2A). London: European Medicines Agency; 1995. Available from: https://www.ema.europa.eu/en/documents/scientific-guideline/international-conference-harmonisation-technical-requirements-registration-pharmaceuticals-human-use-topic-e-2-clinical-safety-data-management-definitions-and-standards-expedited-reporting-step_en.pdf.

20. Teixeira J, Palmisano S. Effects of dynamic field-of-view restriction on cybersickness and presence in HMD-based virtual reality. Virtual Reality. 2021;25:433–445. doi:10.1007/s10055-020-00466-2.

21. Eldridge SM, Chan CL, Campbell MJ, Bond CM, Hopewell S, Thabane L, et al. CONSORT 2010 statement: Extension to randomised pilot and feasibility trials. BMJ. 2016;355: i5239. doi: 10.1136/bmj.i5239.

22. Julious SA. Sample size of 12 per group rule of thumb for a pilot study. Pharm Stat. 2005;4: 287–291. doi: 10.1002/pst.185.

23. Hertzog MA. Considerations in determining sample size for pilot studies. Res Nurs Health. 2008;31: 180–191. doi: 10.1002/nur.20247.

24. Des Jarlais DC, Lyles C, Crepaz N, TREND Group. Improving the reporting quality of nonrandomized evaluations of behavioral and public health interventions: The TREND Statement. Am J Public Health. 2004;94: 361–366. doi: 10.2105/ajph.94.3.361.

25. Humbert A, Kohls E, Baldofski S, Epple C, Rummel-Kluge C. Acceptability, feasibility, and user satisfaction of a virtual reality relaxation intervention in a psychiatric outpatient setting during the COVID-19 pandemic. Front Psychiatry. 2023 Oct 25;14:1271702. doi: 10.3389/fpsyt.2023.1271702.

26. Toussaint A, Hüsing P, Gumz A, Wingenfeld K, Härter M, Schramm E, et al. Sensitivity to change and minimal clinically important difference of the 7-item Generalized Anxiety Disorder Questionnaire (GAD-7). J Affect Disord. 2020;265: 395–401. doi: 10.1016/j.jad.2020.01.032.

27. Lee J, Kim J, Ory MG. The impact of immersive virtual reality meditation for depression and anxiety among inpatients with major depressive and generalized anxiety disorders. Front Psychol. 2024;15: 1471269. doi: 10.3389/fpsyg.2024.1471269.

28. Seong S, Kim H, Cho Y, Kim M-J, Park KR, Choi J, et al. Impact of virtual reality–based biofeedback on sleep quality among individuals with depressive symptoms, anxiety symptoms, or both: 4-week randomized controlled study. J Med Internet Res. 2025;27: e65772. doi: 10.2196/65772.

29. Kim K-Y, Hur M-H, Kim W-J. Effects of virtual reality (VR)-based meditation on sleep quality, stress, and autonomic nervous system balance in nursing students. Healthcare (Basel). 2024;12: 1581. doi: 10.3390/healthcare12161581.

30. de Zambotti M, Barresi G, Colrain IM, Baker FC. When sleep goes virtual: The potential of using virtual reality at bedtime to facilitate sleep. Sleep. 2020;43: zsaa178. doi: 10.1093/sleep/zsaa178.

31. Pardini S, Gabrielli S, Olivetto S, Fusina F, Dianti M, Forti S, et al. Personalized virtual reality compared with guided imagery for enhancing the impact of progressive muscle relaxation training: Pilot randomized controlled trial. JMIR Ment Health. 2024;11: e48649. doi: 10.2196/48649.

32. Garcia LM, Birckhead BJ, Krishnamurthy P, Mackey I, Sackman J, Salmasi V, et al. Three-month follow-up results of a double-blind, randomized placebo-controlled trial of 8-week self-administered at-home behavioral skills-based virtual reality (VR) for chronic low back pain. J Pain. 2022;23: 822–840. doi: 10.1016/j.jpain.2021.12.002.

33. Garcia L, Birckhead B, Krishnamurthy P, Mackey I, Sackman J, Salmasi V, et al. Durability of the treatment effects of an 8-week self-administered home-based virtual reality program for chronic low back pain: 6-month follow-up study of a randomized clinical trial. J Med Internet Res. 2022;24: e37480. doi: 10.2196/37480.

34. Kaussner Y, Kuraszkiewicz AM, Schoch S, Markel P, Hoffmann S, Baur-Streubel R, et al. Treating patients with driving phobia by virtual reality exposure therapy – A pilot study. PLoS One. 2020;15: e0226937. doi: 10.1371/journal.pone.0226937.

